# The Delirium Interview as a new reference standard in studies on delirium assessment tools

**DOI:** 10.1101/2022.12.05.22283028

**Authors:** Fienke L. Ditzel, Arjen J.C. Slooter, Mark van den Boogaard, Michel Boonstra, Timotheus A. van Nesselrooij, Marjan Kromkamp, Monica Pop-Purceleanu, Paul J.T. Rood, Robert Jan. Osse, Carol K. Chan, Alasdair M.J. MacLullich, Zoë Tieges, Karin J. Neufeld, Suzanne C.A. Hut

**Author notes:** **Corresponding author:** Fienke L. Ditzel, MD. Department of Intensive Care Medicine. University Medical Center Utrecht. PO Box 85500 3508 GA Utrecht The Netherlands.

## Abstract

**Background:** The reference standard in studies on delirium assessment tools is usually based on the clinical judgement of only one delirium expert, and may be concise, unstandardized, or not specified at all. This multicenter study investigated the performance of the Delirium Interview, a new reference standard for studies on delirium assessment tools, allowing classification of delirium based on written reports.

**Methods:** We tested the diagnostic accuracy of our standardized Delirium Interview by comparing delirium assessments of the reported results with live assessments. Our reference, the live assessment, was performed by two delirium experts and one well-trained researcher who registered the results. Their delirium assessment was compared with the majority vote of three other independent delirium experts who judged the rapportage of the Delirium Interview. Our total pool consisted of 13 delirium experts with an average of 13±8 years of experience.

**Results:** We included 98 patients (62% male, mean age 69±12 years), of whom 56 (57%) Intensive Care Units (ICU) patients, 22 (39%) patients with a Richmond Agitation Sedation Scale (RASS)<0 and 26 (27%) non-verbal assessments. The overall prevalence of delirium was 28%. The Delirium Interview had a sensitivity of 89% (95% Confidence Interval (CI): 71-98%) and specificity of 82% (95%CI: 71-90%), compared to the diagnosis of an independent panel of two delirium experts and one researcher who examined the patients themselves. Negative and positive predictive values were 95% (95%CI: 86-0.99%) respectively 66% (95%CI: 49-80%). Stratification into ICU and non-ICU patients yielded similar results.

**Conclusion:** The Delirium Interview is a feasible reference method for large study cohorts evaluating delirium assessment tools since experts could assess delirium with high accuracy without seeing the patient at the bedside.

**Impact Statement:** We certify that this work is novel because we propose a new reference standard for delirium assessment tools. It was developed based on an extensive dialogue with multiple delirium experts from different specialties (geriatrics, psychiatry, neurology, neuropsychology, critical care medicine, and nursing), composed from previously validated tests. It is novel in its approach of providing a logistically feasible method that serves as an accurate reference standard.

**Key point box:**

- This paper proposes the Delirium Interview as a new reference for delirium assessment tools used at ICU and non-ICU departments.
- The diagnosis was based on the majority vote of an expert panel of three, accounting for the known disagreement on the diagnosis of delirium.
- Our panel could assess delirium with a high sensitivity of 89% (95% Confidence Interval (CI): 71-98%) and specificity of 82% (95%CI: 71-90%) without being present at the bedside.

**Why does this paper matter?:** Delirium is a major healthcare problem associated with longer hospital duration and increased healthcare costs. Recognition of delirium is essential for the treatment of underlying conditions and optimal communication with affected patients. The Delirium Interview is a reference method standardized for a heterogeneous population, including patients with low consciousness and intubated patients. The methodology is logistically efficient for studies with large cohorts, as we found that delirium experts could diagnose delirium with high accuracy without being present at bedside.

## Introduction

Delirium is a neuropsychiatric syndrome characterized by disturbances of awareness, attention, cognitive functions and behavior.^1^ It occurs in the severely ill, often older patients^2^ and is associated with prolonged hospitalization,^3^ institutionalization, increased costs, long-term cognitive decline and mortality.^2–4^ Recognition of delirium is essential for the treatment of underlying conditions, optimal communication with affected patients and informing family members.^5^ In the previous decade, several delirium assessment tools have been developed.^6^ However, validating these instruments can be subject to various methodological issues.

The reference standard in studies on delirium assessment tools is usually based on the clinical judgement of only one delirium expert and may be concise, unstandardized, or not specified.^6^ The expert relies on diagnostic criteria, most often the Diagnostic and Statistical Manual of Mental Disorders-5 (DSM-5) criteria,^7^ but it is unclear which tests should be used to evaluate these criteria and which cut-off values should be used to interpret these tests.^8^ In the absence of standardized testing, experts often base their diagnosis on unspecified clinical judgement.^9^ Importantly, even if experts have access to exactly the same clinical information, they were found to disagree on delirium classification in 21% of the cases.^10^ To minimize subjectivity and to correct for differences in the level of experience, a panel of experts is needed to classify delirium by majority vote.^9^ These experts must examine the patient simultaneously due to the fluctuating character of delirium and rule out learning effects associated with repeated measurements. However, it appears to be logistically challenging to arrange for two (or more) experienced delirium experts to be present simultaneously, especially when many patients need to be enrolled. Therefore, studies that validate delirium assessment tools are mostly based on small numbers and use a self-selected reference method that is often poorly described.^6,9^

One way to solve the problem of expert availability would be by making video recordings of patients for later classification.^10^ However, video recordings could be burdensome and intimidating for the very ill, vulnerable patient who cannot provide informed consent at the moment of assessment. To overcome challenges pertaining to video-recorded interviews while recognizing the need for an efficient method, we composed the Delirium Interview.

The Delirium Interview is a standardized cognitive test battery capturing all aspects of delirium described by the DSM-5 criteria. It was composed of the input of specialist experts (e.g. geriatricians, psychiatrists) and pre-existing cognitive tests and can be applied to either mechanically ventilated Intensive Care Unit (ICU) or non-ICU patients. This study aimed to investigate to what extent delirium experts can diagnose delirium based on written reports of the Delirium Interview without the need to be present at the bedside.

## Methods

### Setting and study population

This study is part of the DELTA study; a multicenter clinical validation study of DeltaScan for assessing delirium in the ICU and non-ICU wards. The study design was approved by the medical ethical committee of University Medical Center Utrecht, the Netherlands (protocol 17-857), which waived the need for informed consent, and registered at clinicaltrials.gov (NCT03966274). This manuscript adheres to the Standards for Reporting of Diagnostic Accuracy Studies (STARD) guidelines.^11^ This study recruited patients between February 2019 and December 2021 from the University Medical Center Utrecht or the Radboud University Medical Center Nijmegen in the Netherlands. Patients were included when they were admitted to either the ICU (aged 18 years or older), or admitted to the non-ICU ward (aged 60 years or older), and at a consciousness level of a Richmond Agitation Sedation Scale (RASS)^12^ of −2 or higher. Exclusion criteria were: acute brain injury within six weeks prior to the measurement, including post-anoxic encephalopathy and traumatic brain injury; admission because of a primary neurological or neurosurgical disease; a condition hampering delirium assessment (e.g. language barrier or deafness); and pre-existing dementia as documented in the Electronic Health Record (EHR). Patients gave permission before study inclusion and were provided with an opt-out form included in the information letter that was given to them and their family or legal representatives.

### Data collection and study design

The Delirium Interview was conducted by a trained researcher in the presence of two delirium experts: the Live Interview Assessor Panel (LIAP) that we used as a reference. More details on the Delirium Interview are described in Box 1 and Supplement 1-4. After the Delirium Interview, the delirium experts were allowed to ask additional questions to the patient and inspect their EHR. Results of the Delirium Interview were sent out to three other independent delirium experts: the Written Interview Assessor Panel (WIAP). The total pool consisted of 13 delirium experts, with an average of 13 years (standard deviation (SD): 8) of clinical experience. The pool of researchers consisted of six members, trained to conduct the Delirium Interview correctly.

### Statistical analysis

Statistical analyses were performed in Ri386 version 4.0.3. For our main analyses, we tested the performance of the Delirium Interview by comparing the majority vote of the WIAP with the reference: the majority vote of the LIAP (Figure 1). Predictive values, sensitivity, specificity, and overall accuracy were calculated using a 2×2 contingency table and 95% confidence intervals based on 2000 bootstrap samples. We performed stratified analyses according to admittance to the ICU, the ability to verbally communicate, and a decreased level of consciousness indicated by a RASS<0 as compared to RASS=0. Next, interrater reliability for the classification of delirium was calculated with Fleiss’ Kappa within the three raters of a panel, and Cohen’s kappa between two panels. Comparable analyses were performed for the interrater reliability of the delirium probability score (1-10) with the interclass correlation coefficient (ICC, The Oneway Model for randomly unbalanced datasets, including a complete cases analysis since we had more than 5% of missing cases).^13^ More details of the statistical analyses are available in Supplement 5.

**Figure 1.**
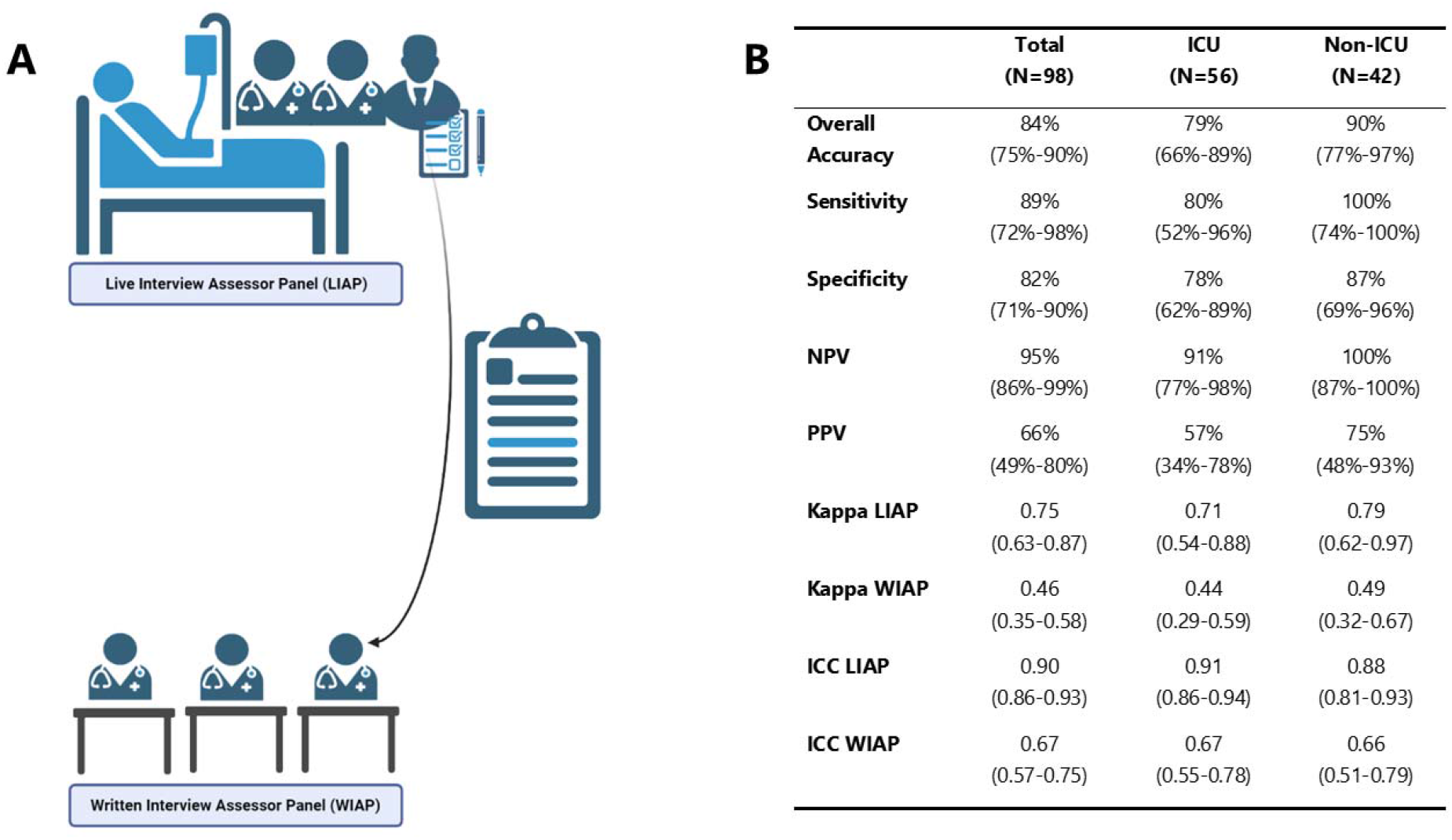
Study design and performance of the Delirium Interview. We tested the diagnostic accuracy of paper-based assessments of the Delirium Interview by comparing assessments of the Live Interview Assessor Panel (LIAP, 2 delirium experts and a researcher) with the Written Interview Assessor Panel (WIAP, 3 different independent delirium experts). Delirium experts were selected out of a pool of 13 and researchers out of a pool of 6. Data are shown as value with 95% Confidence Interval (CI). The delirium probability score included a few missings: 18 (6% of 3×98) within the Live Interview Assessor Panel and 1 (0%) within the Written Interview Assessor Panel. Complete case analysis of the Live Interview Assessor Panel yielded similar results (ICC= 0.89 95%CI: 0.84 - 0.92, N=80).

## Results

### Demographics

In total, 109 patients were assessed by the LIAP, of whom 101 interviews were sent out to the WIAP (for participant flowchart, see Supplement 6). The final cohort consisted of 98 patients, of whom 56 (57%) ICU patients and 42 (43%) non-ICU patients. The cohort included 26 (27%) non-verbal assessments (e.g. due to mechanical ventilation) and 22 (39%) patients had a reduced level of consciousness indicated by a RASS<0. The mean age was 77 (SD: 12) years, 61% (N=60) of the population was male, and the overall prevalence of delirium was 28% (N=27). Patient characteristics are shown in Table 1.

**Table 1.** Patient characteristics

|  | **Total (N=98)** | **ICU (N= 56)** | **Non-ICU (N=42)** |
| --- | --- | --- | --- |
| **Age in years, mean (SD)** | 69 (12) | 63 (11) | 76 (7) |
| **Male sex, n (%)** | 60 (61%) | 34 (61%) | 26 (62%) |
| **Female sex, n (%)** | 38 (39%) | 22 (39%) | 16 (38%) |
| **UMCU, n (%)** | 73 (75%) | 31 (55%) | 42 (100%) |
| **Radboud, UMC n (%)** | 25 (26%) | 25 (45%) | 0 (0%) |
| **Surgical patients, n (%)** | 77 (79%) | 44 (79%) | 33 (79%) |
| **Delirium, n (%)** | 27 (28%) | 15 (27%) | 12 (29%) |
| Hyperactive delirium | 1 (4%) | 1 (7%) | 0 (0%) |
| Hypoactive delirium | 12 (44%) | 7 (47%) | 5 (42%) |
| Mixed delirium | 14 (52%) | 7 (47%) | 7 (58%) |
| **Relevant psychiatric or neurological medical history** |  |  |  |
| Depression, n (%) | 8 (8%) | 6 (11%) | 2 (5%) |
| Alcohol abuse, n (%) | 5 (5%) | 3 (5%) | 3 (5%) |
| CVA, n (%) | 14 (14%) | 6 (11%) | 8 (19%) |
| Parkinson, n (%) | 3 (3%) | 1 (2%) | 2 (5%) |
| Other, n (%) | 8 (8%) | 2 (4%) | 6 (14%) |
| **Relevant medication 2 hours prior to measurement** |  |  |  |
| Antipsychotics, n (%) | 4 (4%) | 2 (36%) | 2 (5%) |
| Alpha-2-antagonists, n (%) | 11 (11%) | 11 (20%) | 0 (0%) |
| Benzodiazepines, n (%) | 2 (2%) | 2 (4%) | 0 (0%) |
| Opioids, n (%) | 19 (19%) | 15 (27%) | 4 (10%) |
| Esketamine, n (%) | 2 (2%) | 2 (4%) | 0 (0%) |
| Propofol, n (%) | 4 (4%) | 4 (7%) | 0 (0%) |
| **Relevant medication 24 hours prior to measurement** |  |  |  |
| Antipsychotics, n (%) | 23 (23%) | 17 (30%) | 6 (14%) |
| Alpha-2-antagonists, n (%) | 17 (17%) | 16 (29%) | 1 (2%) |
| Benzodiazepines, n (%) | 19 (19%) | 12 (21%) | 7 (17%) |
| Opioids, n (%) | 55 (56%) | 33 (59%) | 22 (52%) |
| Esketamine, n (%) | 7 (7%) | 5 (9%) | 2 (5%) |
| Propofol, n (%) | 8 (8%) | 7 (13%) | 1 (2%) |

\*Patients with another psychiatric or neurological history suffered from: epilepsy (n= 1), meningitis (n=2), subdural hematoma 2 months earlier (n=1), intraventricular bleeding (n=1), mild cognitive impairment (n=1) amyotrophic lateral sclerosis (n=1), anxiety disorder (n=1). Registered antipsychotics are Haloperidol, Olanzapine, Quetiapine and Clozapine. Registered alfa-2 antagonists are Clonidine and Dexmedetomidine. Registered benzodiazepines are: Midazolam, Lorazepam, Temazepam, Oxazepam, Diazepam. Registered Opioids are: Morfine, Fentanyl, Remifentanil, Sufentanil, Tramadol, Piritramide, Oxycodon.

### Performance

The overall accuracy of the WIAP was 84% (95%CI: 75-90%) compared to the reference: the LIAP. The Delirium Interview had a sensitivity of 89% (95%CI: 72-98%) and specificity of 82% (95%CI: 71-90%). The negative predictive value (NPV) and positive predictive value (PPV) were 95% (95%CI: 86-99%) and 66% (95%CI: 49-80%), respectively. Overall accuracy was especially high in non-ICU patients (90%, 95%CI: 77-97%) since all delirious patients were recognized by the WIAP (sensitivity of 100%, 95%CI: 74-100%), and there were no false negatives (NPV of 100%, 95%CI 87-100%). Remarkably, sensitivity remained high in patients within non-verbal assessments (90%, 95%CI: 55-100%:) and those with a decreased level of consciousness indicated by RASS<0 (83%, 95%CI: 52-98%), whereas specificity was 64% (95%CI: 35-87%) and 67% (95%CI: 30-93%), respectively. However, we should note that the study was not powered for this stratified analysis. Supplement 7 shows an overview of the performance and the stratified results and Supplement 8 the Details of certainty of Assessor Panel.

### Interrater reliability

Interrater reliability was substantial within the LIAP with a kappa coefficient of 0.75 (95%CI: 0.63-0.87) and moderate in the WIAP (0.46, 95%CI: 0.35-0.58).^14^ Agreement was higher in the delirium probability score with an excellent Intraclass Correlation (ICC) within LIAP (0.90 95%CI: 0.86-0.93) and moderate ICC in the WIAP (0.67, 95%CI: 0.57-0.75).^15^ The delirium probability score included a few missings: 18 (6%) within the Live Interview Assessor Panel and 1 (0%) within the Written Interview Assessor Panel. Complete case analysis of the Live Interview Assessor Panel yielded similar results (ICC= 0.89 95%CI: 0.84 - 0.92, N=80).

## Discussion

In summary, this prospective multicenter study assessed the diagnostic accuracy of the Delirium Interview, a proposed reference standard for studies on delirium assessment tools for ICU and non-ICU settings. We compared paper-based assessments of the Delirium Interview with live assessments of patients and observed a sensitivity of 88% (95%CI: 72-98%) and specificity of 82% (95%CI: 71-90%).

Our results show that this method is accurate and provides important logistical benefits, as it allows the experts to classify delirium at a suitable moment instead of performing a time-consuming bedside assessment. Therefore, this method seems feasible in studies on delirium assessment tools, especially when a large number of patients needs to be enrolled.

Our moderate^14^ interrater reliability of the WIAP was similar to video assessments of patients (kappa=0.61).^10^ As expected, the LIAP showed a higher kappa of 0.78, which was comparable with studies investigating interrater reliability of brief delirium monitoring tools in live assessments.^16^These results confirm that although experts may disagree on the classification of delirium, relying on the majority vote of delirium experts yields high diagnostic accuracy. Interrater reliability of our delirium probability scale showed less disagreement than the final diagnosis (delirium/no delirium). This is in accordance with the concept to regard delirium rather as a continuous spectrum instead of a dichotomous entity.^17–19^ We recommend that future studies on delirium assessment tools always use the classification of at least three delirium experts and include a delirium probability scale to overcome this problem. To minimize subjectivity, it would be best to randomly pair three experts from a considerably large panel with experience in various clinical settings.

The following issues were encountered based on feedback from our Interview Assessor Panels on the Delirium Interview. First of all, we received feedback that the Month Of The Year Backwards (MOTYB) was one of the most informative and reliable components of the Delirium Interview. A study focusing on the accuracy of attention tests in delirium concluded that no test could become a single-item screening tool, but that the MOTYB showed the best balance between sensitivity and specificity.^20^ Secondly, it appears crucial to report the reason for a missing item of the Delirium Interview. One study showed that half of the patients who refused to answer questions on an attention test were assessed as delirious by a second21examiner. Thirdly, assessors acknowledged having difficulties in diagnosing subsyndromal delirium. In the medical setting, resolving delirium in an improving patient has less diagnostic value, but in research, there is a definite need to define this group of patients. Some experts mentioned that the Delirium Interview could be improved by adding the educational level of the patient, although others disagreed and argued that anyone without an altered mental state should have been able to perform all items of the test, independent of their prior education. Finally, the ‘narrative’ in which the researcher could include subjective elements in the patient report appeared to be a section of great value to the Delirium Interview. To pursue a good narrative, it is crucial that independent researchers are appropriately trained, have intervision sessions and are supervised in the beginning as well as regularly thereafter.

The strengths of this study are the multicenter setting on ICU and non-ICU departments, the various group of delirium experts and the expertise used to compose the Delirium Interview. A limitation of the Delirium Interview is that we did not include an objective measurement of pre-existent cognitive status. Since dementia affects all the selected attentional and cognitive tests,^22^ we chose to exclude patients with cognitive impairment based on the medical history reported in the EHR. Further research could explore a reference standard for delirium superimposed on dementia.^23^ Performance was not majorly influenced by difficult subgroups since sensitivity remained high, and specificity sufficient in patients unable to communicate verbally (e.g., mechanically ventilated patients) and in those with a low level of consciousness. However, our sample was unpowered for such stratified analysis, thus we recommend future studies to focus on the isolated performance of these patients. Furthermore, future studies may benefit from exploring delirium severity, for example, by ranking the various items of the Delirium Interview.

## Conclusion

Delirium experts could assess delirium with high accuracy based on written reports and responses to the Delirium Interview without being present at the bedside, which makes this method feasible to study large cohorts. We believe that the Delirium Interview may serve as an appropriate reference standard for the diagnosis of delirium for both ICU and non-ICU patients when used in studies on the evaluation of delirium assessment tools.

## Supporting information

Supplemental Material Delirium Interview

## Data Availability

All data produced in the present study are available upon reasonable request to the authors

## Conflict of interests

None of the other authors reports any conflicts of interest.

## Author Contributions

All authors confirmed approval for the manuscript to be published.

<u>Fienke L. Ditzel, MD:</u> This author contributed to data collection, data preparation, analysis and preparation of the manuscript.

<u>Arjen J.C. Slooter, MD PhD:</u> This author contributed to the study concept and design, composing the Delirium Interview, data collection and revision of the manuscript.

<u>Mark van den Boogaard, RN PhD:</u> This author contributed to the study concept, composing the Delirium Interview, data collection and revision of the manuscript

<u>Michel Boonstra, MSc:</u> This author contributed to the study concept and design, composing the Delirium Interview, data collection, data preparation and revision of the manuscript.

<u>Tim van Nesselrooij</u>: This author contributed to data collection.

<u>Marjan Kromkamp, MD PhD</u>: This author contributed to data collection.

<u>Monica Pop-Purceleanu, MD PhD</u>: This author contributed to data collection and revision of the manuscript.

<u>Paul J.T. Rood, RN PhD:</u> This author contributed to data collection.

<u>Robert Jan Osse MD PhD:</u> This author contributed to data collection.

<u>Carol K. Chan, MBBCh, MSc</u>: This author contributed to revision of the manuscript. <u>Alasdair M.J. MacLullich, MRCP PhD:</u> This author contributed to the study concept, composing the Delirium Interview and revision of the manuscript.

<u>Zoë Tieges, PhD:</u> This author contributed to the study concept, composing the Delirium Interview and revision of the manuscript.

<u>Karin J. Neufeld, MPH MD:</u> This author contributed to the study concept, composing the Delirium Interview and revision of the manuscript.

<u>Suzanne C.A. Hut, PhD:</u> This author contributed to the study concept and design, composing the Delirium Interview, data collection, data preparation, analysis and revision of the manuscript.

We would like to thank A. Vondeling MD, C. H. Röder MD PhD, J.W.M. Krulder MD, R. Faaij MD PhD, A. Kamper MD, A. de Jonghe MD PhD, P. Bouvy, MD PhD, E. van Dellen MD PhD, K. Milisen RN PhD, B. van Munster MD PhD and especially M. Coesmans for their share in the paper-based assessments of the Delirium Interview and K. Luijken PhD for help with the analysis.

## Sponsor’s Role

This work was supported by European Union Horizon 2020 [grant number 820555]. The sponsor had no role in the study design, data collection, data analysis, data interpretation, writing of the report, or the decision to submit for publication.

---

**Box 1.**
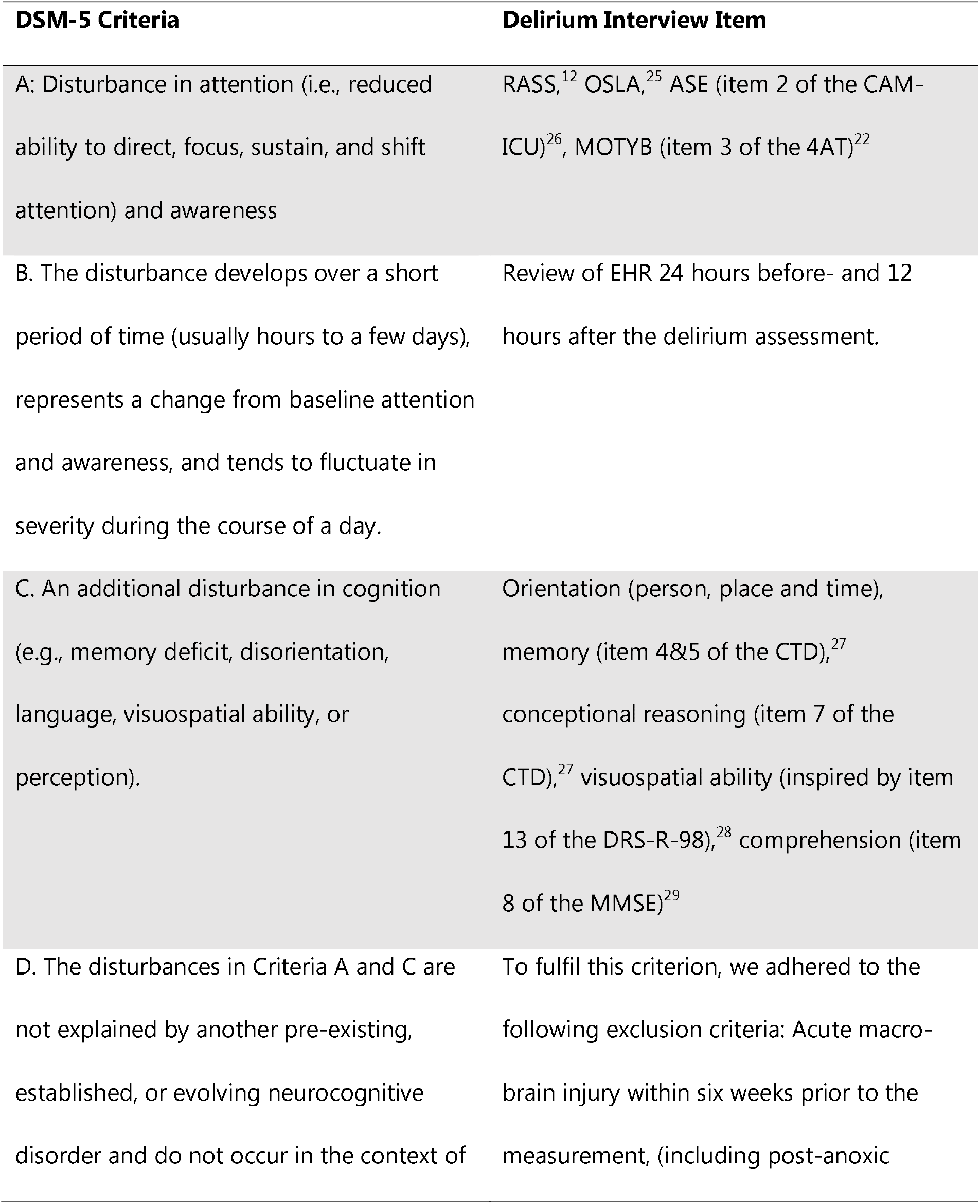
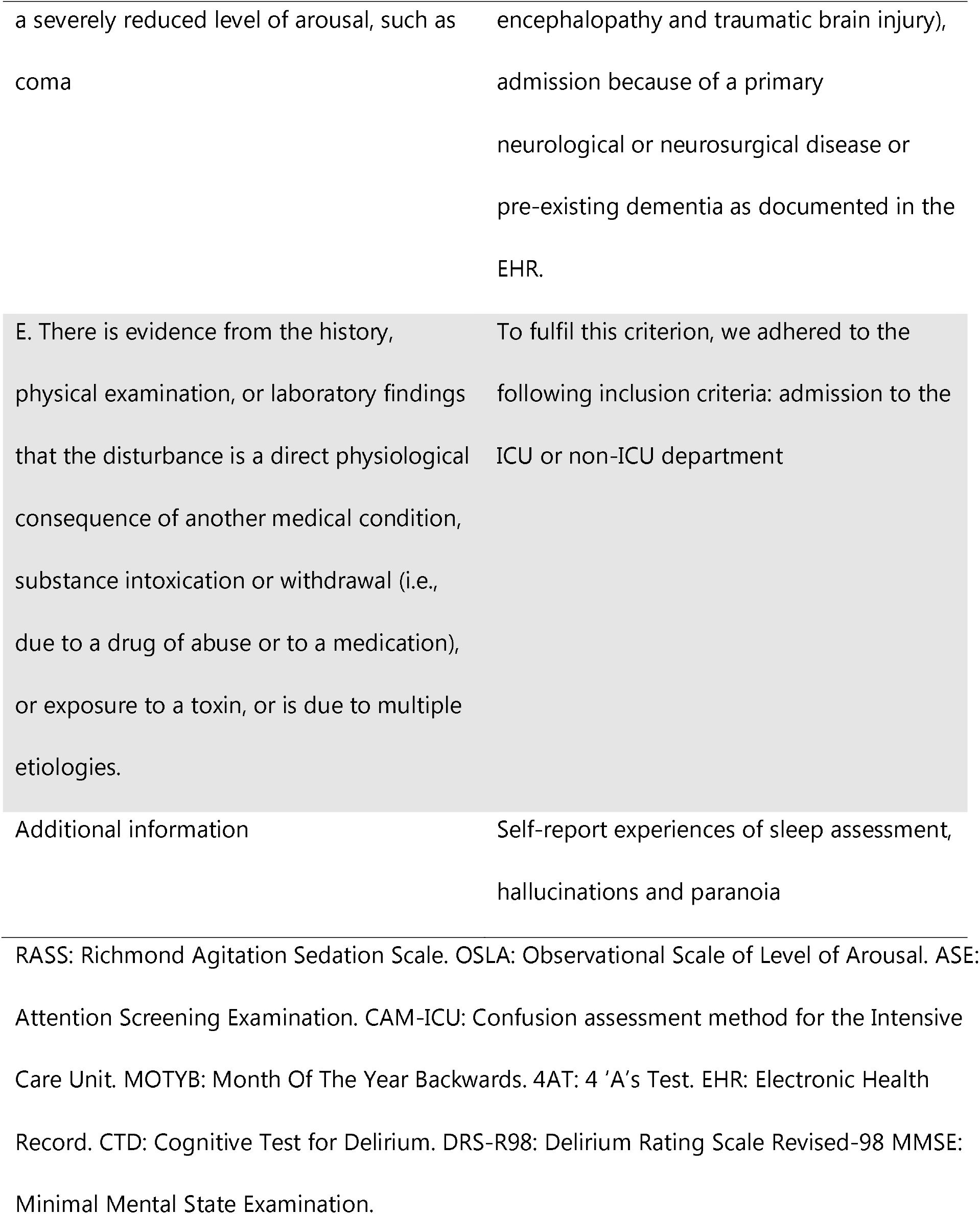
The DSM-5 criteria and the items of the Delirium Interview.

## Notes

### Competing Interest Statement

The authors have declared no competing interest.

### Clinical Protocols

https://clinicaltrials.gov/ct2/show/NCT03966274?cond=deltascan&draw=2&rank=3

### Funding Statement

This work was funded by European Union Horizon 2020 [grant number 820555].

### Author Declarations

Ethics committee of University Medical Center Utrecht, the Netherlands (protocol 17-857) waived ethical approval for this work

