## Supplemental Material Delirium Interview for "The Delirium Interview as a new reference standard in studies on delirium assessment tools"

### **Supplement 1: The development of the Delirium Interview**

The interview was developed based on an extensive dialogue with multiple delirium experts from different specialties (geriatrics, psychiatry, neurology, neuropsychology, critical care medicine, and nursing). The main goals were to follow the DSM-5 criteria for delirium and make a non-verbal variant for e.g. ventilated patients. To accomplish the latter, each item was presented as a multiple-choice question on an A4-sized card, with a standard question formulation on the back of the card visible to the researcher, to ensure standardized test administration. The researcher systematically pointed out all the options and ventilated patients nodded or blinked when the researcher pointed at the correct answer.

The Delirium Interview is a combination of different previously validated tests, including the Richmond Agitation Sedation Scale (RASS),^12^ the Observational Scale of Level of Arousal (OSLA),^30^ the 4 ‘A’s Test (4AT),^22^ Confusion Assessment Method for the Intensive Care Unit (CAM-ICU),^31^ the Cognitive Test for Delirium (CTD)^27^ and the Schedules for Clinical Assessment in Neuropsychiatry (SCAN).^32^ After completing the assessment, the researcher was prompted to write a short narrative regarding the general impression of the patient including his/her reaction upon arrival, extent of eye contact, reaction speed, (abnormal) motoric movement and whether communicative behavior was appropriate. The narrative also provided the option to inform the Written Interview Assessor Panel (WIAP) of the opinion of nurses and family members. As this information was not always provided, we made this topic facultative and unstandardized. The median duration of the Delirium Interview was 10 minutes (IQR: 5-15 minutes). The full interview and its Dutch translation as used in the current study, are presented in Supplement 2.

All answers to the items were summarized systematically and consistently in the Delirium Score Card, including notes on possible mistakes the patients made. We included surgery type, neurological and psychiatric history, presence of visual or hearing aids, and medical information from the EHR in the 24 hours prior to the interview and 12 hours after the interview. The Delirium Score Card was developed in a manner that should optimize efficient scoring of the DSM-5 criteria A, B and C, therefore aiming to allow a delirium expert to reach a clinical diagnosis without having been present at the bedside (Supplement 3 for filled examples of the Delirium Score Card).

### **Supplement 2: The Delirium Interview**

**Delirium Interview**

**1.1 Arousal - RASS score**

-3	Moderate sedation: Movement or eye opening to voice (but no eye contact)

-2	Light sedation: Briefly awakes to voice (eye opening & contact < 10 seconds)

-1	Drowsy: Not fully alert but has sustained awakening to voice (eye contact ≥ 10 seconds)

0	Alert and calm

+1 Restless: Anxious, apprehensive, but movements not aggressive

+2 Agitated: Frequent nonpurposeful movement, fights ventilator

+3 Very agitated: Pulls or removes tube(s) or catheter(s), aggressive

**1.2 Arousal – OSLA**

| **Eye Opening** | |  | **Eye Contact** | |
| --- | --- | --- | --- | --- |
| *Score* | *Description* |  | *Score* | *Description* |
| **0** | Are open on arrival and remain so, under patient’s control, outlasts stimulus |  | **0** | Spontaneously makes and holds eye contact appropriately |
| **1**-1 | Are open on arrival but close if stimulus removed |  | **1**-1 | Drowsy and makes eye contact to command but can’t hold it for very long |
| **1**-2 | Open to voice but then outlast stimulus |  | **1**-2 | Alert but eyes wandering, some appropriate eye contact |
| **2** | Open to voice but close if stimulus removed |  | **2**-1 | Alert but eyes wandering, little or no appropriate eye contact |
| **3** | Open to gentle physical stimulation (squeezing hand, gently shaking shoulder) |  | **2**-2 | Drowsy but makes brief eye contact |
| **4** | Open to pain only |  | **3** | Eyes will/are open but no eye contact |
| **5** | No eye opening |  |  | |
| **Posture (NB take into account weakness due to stroke or neurological disease, etc.)** | |  | **Movement** | |
| *Score* | *Description* |  | *Score* | *Description* |
| **0** | Sitting out in chair or up in bed, holding appropriate posture |  | **0** | Moves spontaneously and purposefully with no restless or agitated movements |
| **1** | Slumped in chair or bed but attempts to sit upright and sustain posture on request |  | **1**-1 | Occasional or mild restless or fidgety movements, no aggressive or vigorous movements |
| **2** | Slumped in chair or bed and unable to sustain posture |  | **1**-2 | Reduced frequency of movement, mildly slowed up |
| **3** | Lying in bed and unable or no response to request to sustain posture |  | **2**-1 | Frequent restless or fidgety movements, no aggressive or vigorous movements |
|  |  |  | **2**-2 | Moderately reduced frequency and speed of movement, interfering with assessment or self-care |
|  |  |  | **3** | Aggressive or vigorous, recent pulling out of lines |
|  |  |  | **4**-1 | Overtly combative, violent |
|  |  |  | **4**-2 | Severely reduced frequency and speed of movement, few spontaneous movements |

**1.3 Communication barriers**

Able to communicate verbally: Yes/No

If no, specify reason (e.g. intubation, mechanical ventilation): …………………………

Able to nod/shake head: Yes/No

If no, able to communicate via hand squeezing? Yes/No

If no, able to communicate via eye blinks? Yes/No

Eye glasses or hearing aid? Yes/No

**2. Orientation**

a. **Am I right that your age is [*offer options below*]** ?	□ No answer

□ 2 years younger	□ 30 years younger

□ Correct age	□ 10 years younger

b. **Is it correct that you were born in [*offer options below*] ?**

□ No answer

□ Wrong month(+1)	□ Correct date of birth

□ Wrong year(-10)	□ Wrong day(+1)

**I’m now going to show you some text on this card. Can you read this well? Okay, great.**

*(if patient is not able to read or does not focus on the card, then read each option aloud with an interval of 3 seconds and ask to say yes/no (or alternative communication) upon right/wrong answers)*

**c. Could you tell me what year it currently is?**

□ No answer

□ 2 years earlier	□ 30 years earlier

□ correct year	□ 10 years earlier

d. **And what month is it?**

□ No answer

□ 4 months earlier	□ 2 months later

□ correct month	□ 4 months later

e. **What time of day is it currently?**

□ No answer

□ Morning	□ Night

□ Evening	□ Afternoon

f. **Where are you at this moment?**

□ No answer

□ Doctor's office	□ Home

□ Airport	□ Hospital

**3. Attention**

**I’m going to give you my hand, could you please squeeze it once (*alternative: could you please nod your head for me once*)? Thank you. This is important for the next test, because you will squeeze/nod some more. I am going to read to you a series of 10 letters. Whenever you hear the letter ‘A,’ squeeze my hand/nod your head. When you hear any other letters, you don’t have to squeeze. Here we go.**

S / A / V / E / A / H / A / A / R / T	□ No response

Number correct: …. / ….

Speed of task completion: swiftly / normal / slow

b. **Here is a list of the months of the year. I would like to ask you to point to the months of the year - or say them aloud if you prefer - but in reversed order. So you can start with ‘December’ and go all the way back.**	□ No response

…………………………………………………………………………………………………………….	…………………………………………………………………………………………………………….

Number correct: …. / 12

Speed of task completion: Swiftly / normal / slow

**4. Short term memory**

*First assess whether the patients is able to focus on card by moving the card from the left to the right in front of them and check if they follow the card with their eyes.*

□ Patient unable to complete

**Now I am going to show you three pictures of common objects. Watch carefully and try to remember each picture, because later I will ask you something about them again.** *Name each object as you point to it. Show each picture for 3 seconds.* *Circle form used. In case the patient cannot see well, you can ask them to remember the words you say aloud.*

□ Form A: table car hammer	□ Form B: dog knife pants

**Thank you very much. In a few minutes I will ask you something about them again, so please try to remember them all. We will first do something else.**

**5. Visuospatial ability	□** Patient unable to complete

a**. Could you please point to the door of this room?**

□ Incorrect	□ Correct	□ No answer

b. **And what is closer to you – is that me** (*point to self*) **or the door** (*point to the door*)**?**

□ Incorrect	□ Correct	□ No answer

c. **Here is a card with some figures. Could you please point to the square?**

□ Incorrect	□ Correct	□ No answer

d. **And could you now point to the largest circle?**

□ Incorrect	□ Correct	□ No answer

**6. Conceptual reasoning**

□ Patient unable to complete

**Which one of these does not belong to the same group as the other three? Point to the correct answer.** *Read each answer as you point to it, and circle choice. Circle form used.*

□ Form A	□ Form B

a. Bus Airplane Bicycle Apple:	a. Table Couch Desk Goat:

□ Incorrect	□ Correct	□ No answer	□ Incorrect	□ Correct	□ No answer

b. Arm House Foot Nose:	b. Dress Corn Shirt Shoes:

□ Incorrect	□ Correct	□ No answer	□ Incorrect	□ Correct	□ No answer

**7. Short term memory II**

□ Patient unable to complete

**Earlier I asked you to remember the three pictures I showed you. Now I am going to show you some more pictures. Some you have just seen but others will be shown for the first time. Let me know whether or not you saw the picture before by saying ‘yes’/nodding your head yes** *(demonstrate)* **or saying ‘no’/nodding your head no** *(demonstrate)***.**

**[***show picture***] Have you seen this picture before?** *[Etc.]* **Well done, thank you.**

Form A	Form B

a. Car (yes)	□ Yes	□ No	□ No answer	a. Fork (no)	□ Yes	□ No	□ No answer

b. Glass (no)	□ Yes	□ No	□ No answer	b. Knife (yes)	□ Yes	□ No	□ No answer

c. Truck (no)	□ Yes	□ No	□ No answer	c. Dog (yes)	□ Yes	□ No	□ No answer

d. Table (yes)	□ Yes	□ No	□ No answer	d. Sock (no)	□ Yes	□ No	□ No answer

e. Hammer (yes) □ Yes	□ No	□ No answer	e. Pants (yes)	□ Yes	□ No	□ No answer

**8. Comprehension**

a. **Please read this and do what it says**.

[*Show examinee the page with CLOSE YOUR EYES*]

□ Incorrect	□ Correct	□ No answer

b. **Listen carefully because I am going to ask you to do three things. You may first listen and when I’m done, you may do what I asked of you. Here we go: Could you please raise your eyebrows, stick out your tongue, and shake your head no?** *(If patient is unable to perform these actions, make up alternatives, such as blinking/squeezing/nodding)*

□ Incorrect	□ Correct	□ No answer

**9. Self-report experiences of sleep assessment, hallucinations and paranoia**

a. **Did you sleep well last night?**

□ Yes	□ No	□ No answer

b. *[If no],* **What happened? Were you able to fall back asleep easily after being awoken?**

□ Yes	□ No	□ No answer

c. **Do you feel that the staff members are taking good care of you?**

□ Yes	□ No	□ No answer

d. *[If no],* **Have you felt safe here in the last day?**

□ Yes	□ No	□ No answer

e. **Have you seen or heard anything in the hospital since you’ve been here, that others may not see, for example spots or tiny bugs?**

□ Yes	□ No	□ No answer

f. *[If yes],* **Have you experienced anything like that in the last day?**

□ Yes	□ No	□ No answer

g. **And since being in the hospital, have you had any unusual sensations such as objects changing in size, shape or color?**

□ Yes	□ No	□ No answer

h. *[If yes],* **Have you experienced anything like that in the last day?**

□ Yes	□ No	□ No answer

i. **At the moment, are you able to think clearly? And do you feel like your thinking is the same as it was before you got sick and had to come to the hospital this time?**

□ Yes	□ No	□ No answer

**11. Time completed (24h scale): _____:_______**

*Hour	Minute*

**12. Narrative by Assessor**:

*Describe your general impression of the patient; how did the patient react to you upon arrival; was there any eye contact; did the patient appear drowsy or slow in reacting; did you observe anything abnormal about the motoric movements of the patient; was (communicative) behaviour appropriate; or did you notice anything else that may be important (e.g. in sleep assessment)? Specific examples are helpful.*

______________________________________________________________________________________________________________________________________________________________________________________________________________________________________________________________________________________________________________________________________________________________________________________________________________________________________________________________________________________________________________________________________________________________________________________________________________________________________________________________________________________________________________________________________________________________________________________________________________________________________________________________________________________________________________________________________________________________________________________________________________________________________________________________________________

### **Supplement 3: Overview of known Delirium monitoring scales and the incorporation in the Delirium Interview**

| 4-AT | Delirium Interview |
| --- | --- |
| Alertness | A more extended reflection of this item is represented in the RASS (item 1.1) and OSLA (item 1.2) |
| Abbreviated mental 4 | Item 2 |
| MOTYB | Item 3.a |
| Acute change in alertness and cognition | Review of EHR 24 hours before- and 12 hours after the delirium assessment |
| **CAM-ICU** |  |
| Change or fluctuating mental status | Review of EHR 24 hours before- and 12 hours after the delirium assessment |
| Inattention / SAVEAHEART | Item 3.b |
| RASS | Item 1.1 |
| Disorganized thinking | A more extended reflection of this item is represented in item 5 (visuospatial function), item 6 (conceptual reasoning), item 8 (comprehension) |
| **DRSR-R-98** |  |
| 1 Sleep-wake cycle disturbance | Item 9.a, 9.b |
| 2 Perception and hallucinations | Item 9.e, 9.f, 9.g, 9.h |
| 3 Delusions | Item 9.c, 9.d |
| 4 Lability of effect | Item 9.c, 9.d |
| 5 Language | Present throughout the interview in the verbal variant |
| 6 Though process abnormalities | Item 5 (visuospatial function) Item 6 (conceptual reasoning) |
| 7 Motor agitation | Item 1.2 |
| 8 Motor retardation | Item 1.2 |
| 9 Orientation | Item 2 |
| 10 Attention | A more extended reflection of this item is represented in item 3 (SAVEAHEART/ASE and MOTYB) |
| 11 short-term memory | Item 4 |
| 12 Longterm memory | Item 7 |
| 13 Temporal onset of symptoms | Review of EHR 24 hours before- and 12 hours after the delirium assessment |
| 14 fluctuation in symptom severity | Review of EHR 24 hours before- and 12 hours after the delirium assessment |
| 15 Physical disorder | Inclusion criteria |
| **DOSS** |  |
| 1 Dozes during conversation or activities | Item 1.2 OSLA |
| 2 Is easily distracted by stimuli from the environment | Item 3 attention (SAVEHEART/ASE) |
| 3 Maintains attention to conversation or action | Item 3 attention (SAVEHEART/ASE) |
| 4 Does not finish question or answer | Item 3 attention (SAVEHEART/ASE) |
| 5 Gives answers that do not fit the question | Item 5 Visuospatial ability, Item 6 conceptual reasoning, Item 8 comprehension |
| 6 Reacts slowly to instructions | Duration of the interview |
| 7 Thinks to be somewhere else | Item 2.f |
| 8 Knows which part of the day it is | Item 2.e |
| 9 Remembers recent event | Item 2 Orientation / Item 7 memory |
| 10 Is picking, disorderly, restless | RASS / item 1.2 OSLA |
| 11 Pulls IV tubes, feeding tubes, catheters etc | 1.2 OSLA |
| 12 Is easily or suddenly emotional | Item 9.c, 9.d,9.i |
| 13 Sees / hears things which are not there | Item 9.e-h |
| **ICDSC** |  |
| 1 Altered level of consciousness (SAS or RASS) | Item 1.1 |
| 2 Inattention | This observation will be tested in item 3 (SAVEHEART/ASE and MOTYB) and item 8 comprehension |
| 3 Orientation | Item 2 |
| 4 Hallucinations | Item 9.e-h |
| 5 Psychomotor agitation/retardation | Item 1.2 Osla |
| 6 Inappropriate speech or mood | Mood was tested in item 9.c,d paranoia. Speech could only be tested in the verbal variant of the test. |
| 7 Sleep/wake cycle disturbance | Item 9.a,b |
| 8 Symptom fluctuation | Review of EHR 24 hours before- and 12 hours after the delirium assessment |

### **Supplement 4: Statistical analysis**

Statistical analyses were performed in R i386 version 4.0.3. Descriptive data were presented as frequencies with percentages for categorical data, and either as mean with standard deviation (SD), or median with interquartile range (IQR) for continuous data, depending on the distribution. Data distribution was tested for normality by visually inspecting histograms and normal quantile-quantile plots. For our main analyses, we tested the performance of the Delirium Interview by comparing the majority vote of the WIAP with the reference: the majority vote of the LIAP. Both the LIAP and the WIAP were asked to classify the state of the patient as “no delirium” or “delirium”, to record their motor subtype (hypoactive- hyperactive or mixed delirium) and to express the level of certainty as a number between 1 (being certain: no delirium) and 10 (being certain: delirium), individually and independently of each other. The final diagnoses (“delirium” or “no delirium”) were determined by the majority vote. When in disagreement on the motor subtype, ‘mixed’ was assigned. Predictive values, sensitivity, specificity, and overall accuracy were calculated using a 2x2 contingency table and 95% confidence intervals based on 2000 bootstrap samples. In addition, we performed stratified analyses according to admittance to the ICU, the ability to verbally communicate, and decreased level of consciousness indicated by a RASS<0 compared to RASS=0. Next, interrater reliability for the classification of delirium was calculated with Fleiss’ Kappa within the three raters of a panel, and Cohen’s kappa between two panels. Comparable analyses were performed for the interrater reliability of the delirium probability score (1-10) with the interclass correlation coefficient (ICC, The Oneway Model for randomly unbalanced datasets).^13^

### **Supplement 5: Figure S1 Participant flowchart**

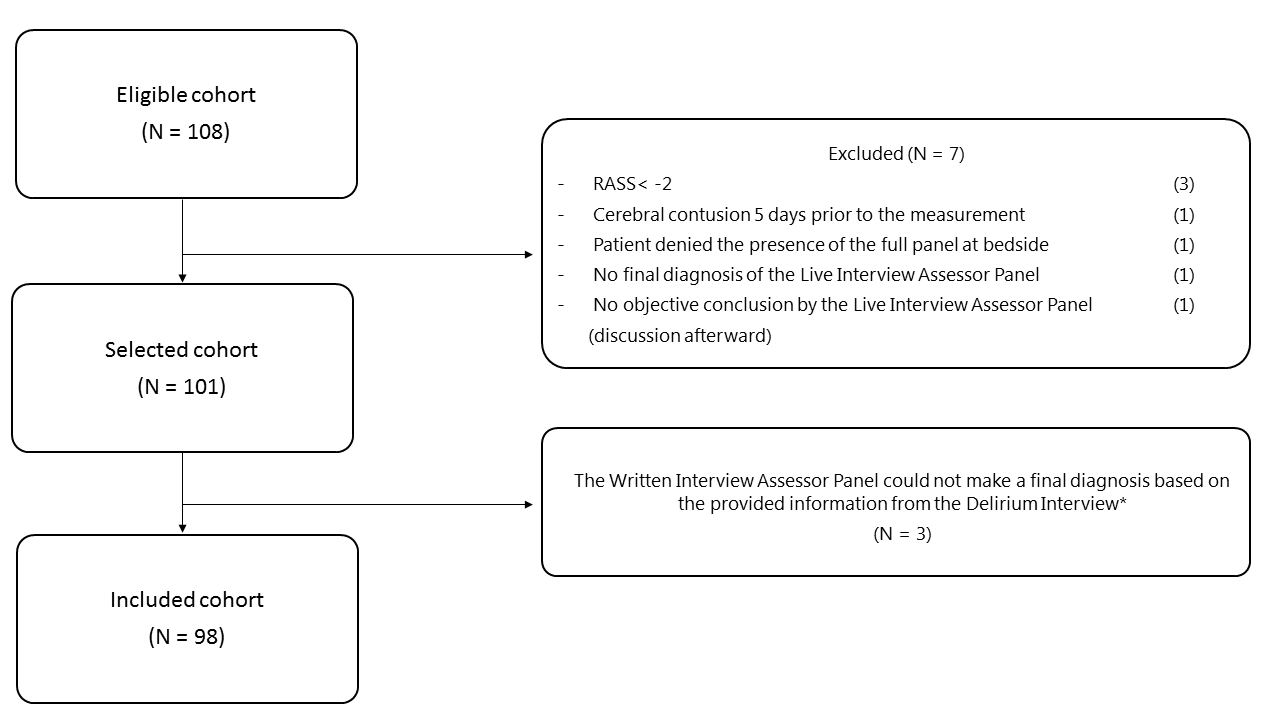
*The Live Interview Assessor Panel was unable to diagnose three patients based on the Delirium Interview. In these cases, the interviews lacked information because too many questions were skipped (e.g. due to interrupting medical staff or because the patient did not answer).

### **Supplement 6: Stratified performance of the Delirium Interview**

|  | **Total**  **(N=98)** | **ICU**  **(N=56)** | **Non-ICU (N=42)** | **Verbal**  **(N=74)** | **Non-verbal (N=24)** | **RASS=0**  **(N=75)** | **RASS<0 (N=21)** |
| --- | --- | --- | --- | --- | --- | --- | --- |
| **Overall Accuracy** | 84%  (75%-90%) | 79%  (66%-89%) | 90%  (77%-97%) | 86%  (77%-93%) | 75%  (53%-90%) | 76%  (52%-92%) | 76%  (53%-92%) |
| **Sensitivity** | 89%  (72%-98%) | 80%  (52%-96%) | 100%  (74%-100%) | 88%  (64%-99%) | 90%  (55%-100%) | 92%  (72%-92%) | 83%  (52%-98%) |
| **Specificity** | 82%  (71%-90%) | 78%  (62%-89%) | 87%  (69%-96%) | 86%  (74%-94%) | 64%  (35%-87%) | 84%  (30%-93%) | 67%  (30%-93%) |
| **NPV** | 95%  (86%-99%) | 91%  (77%-98%) | 100%  (87%-100%) | 96%  (87%-100%) | 90%  (55%-100%) | 98%  (90%-100%) | 75%  (35%-97%) |
| **PPV** | 66%  (49%-80%) | 57%  (34%-78%) | 75%  (48%-93%) | 65%  (43%-84%) | 64%  (35%-87%) | 55%  (32%-76%) | 77%  (46%-95%) |

RASS = Richmond Agitation Sedation Scale. Data are shown as value with 95% Confidence Interval (CI). There were 2 patients with a RASS>0 (both RASS+1), both delirious and classified correctly (accuracy =100%). Overall accuracy was defined as (the true positives + the true negatives)/all cases.

### **Supplement 7: Uncertainty of delirium**

At the end of the Interview the delirium experts of the WIAP were asked to assess a patient as “delirium”, “uncertain” or “no delirium”. Thereafter we asked them to make a forced decision: “delirium” or “no delirium”. The forced decision was used to calculate the majority vote of the panel. Since delirium status was assessed by the WIAP consisting of three delirium experts, a total of 294 (3 times 98) written assessments were performed. In 80% (n=236) of these assessments, the WIAP noted they were certain about their diagnosis. Reasons for being uncertain before giving the final diagnosis were: limited information (n=24, 8%), subsyndromal delirium (n=29, 10%), abnormal behavior possibly caused by medication (n=3, 1%), abnormal behavior possibly pre-existent (n=1, 0%) or unspecified (n=1, 0%).
